# Spine-Related Conditions in MEPS 2020: Association Between Care Fragmentation and Outpatient MRI/CT Use and Radiology Expenditures

**DOI:** 10.1101/2025.08.24.25334318

**Authors:** Andrew Bouras

## Abstract

Outpatient advanced imaging is central to the evaluation of spine-related conditions, yet concerns persist about coordination and potential overuse. We assessed whether higher care fragmentation is associated with increased MRI/CT use and radiology expenditures among U.S. adults with spine-related conditions using 2020 Medical Expenditure Panel Survey (MEPS) data. A spine cohort was identified from ICD-10-CM and CCSR codes; care fragmentation was categorized using fixed cutpoints on full-year utilization. Outcomes included MRI/CT-only imaging in office-based care and radiology expenditures analyzed with survey-weighted regression models. Compared with Low fragmentation, Medium and High fragmentation were associated with higher odds of advanced imaging and higher odds of any radiology spending; conditional spending was directionally higher for High vs Low but imprecise. Effects appeared strongest among privately insured. Findings suggest opportunities to improve coordination and value in ambulatory spine care.

## 1 Introduction

Spine-related conditions are common and often evaluated with advanced imaging in ambulatory care. While MRI and CT can be pivotal for diagnosis, concerns persist regarding coordination and potential overuse when imaging occurs outside of red-flag indications. Appropriateness frameworks emphasize restraint for uncomplicated back pain and timely imaging for red-flags [2, 3, 4, 5]. We examine whether greater care fragmentation is associated with increased outpatient MRI/CT use and higher radiology expenditures in a nationally representative sample of U.S. adults with spine-related conditions.

We used 2020 Medical Expenditure Panel Survey (MEPS) data to build a person-year dataset, identified a spine cohort using diagnosis codes, and categorized care fragmentation using fixed, reproducible cutpoints on annual utilization [6]. Outcomes included MRI/CT-only imaging in office-based care and radiology expenditures, analyzed with survey-weighted models. Our goal is to provide policy-relevant, population-level evidence to inform efforts to improve coordination and value in spine imaging.

## 2 Methods

### 2.1 Data source and cohort

We used MEPS 2020: Full Year Consolidated (HC-224), Office-Based Medical Provider Visits (HC-220G), and Medical Conditions (HC-222) [6]. Adults (AGELAST ≥ 18) with non-missing survey design variables were included. The spine cohort was defined by ICD-10-CM (M48, M50–M54) and/or CCSR categories for spinal stenosis, intervertebral disc disorders, spondylosis, or low back pain.

### 2.2 Fragmentation exposure

Care fragmentation (`fragmentation_score_cat`) was categorized on fixed, reproducible cutpoints using annual utilization variables (`OBTOTV20, ERTOT20, IPDIS20`).

**Table 1:** Fragmentation category definitions.

| Level | Definition |
| --- | --- |
| None | OBTOTV20=0 & ERTOT20=0 & IPDIS20=0 |
| Low | IPDIS20=0 & ERTOT20=0 & OBTOTV20 in [1,4] |
| Medium | IPDIS20=0 & (OBTOTV20 in [5,9] or ERTOT20≥1) |
| High | IPDIS20≥1 or OBTOTV20≥10 or (ERTOT20≥1 & OBTOTV20≥5) |

### 2.3 Outcomes and covariates

Primary outcome: any MRI/CT-only imaging in office-based (OB) care in 2020. Advanced imaging flags were derived from HC-220G service indicators (`MRI_M18, SONOGRAM_M18, XRAYS_M18, MAMMOG_M18`). Secondary outcomes: any advanced imaging (MRI/CT±ultrasound), radiology expenditures (two-part: any spend; positive spend), and count of imaging-positive OB visits. Covariates: age group, sex, race/ethnicity, poverty status, insurance type, total OB visits (`OBTOTV20`), multimorbidity (`mcc_2plus`), and design variables (`PERWT20F, VARSTR, VARPSU`).

### 2.4 Statistical analysis

Survey-weighted logistic regression was used for binary outcomes; quasi-Poisson for counts; two-part model for expenditures (quasibinomial for any spend; quasi-Poisson with log link for positive spend). Survey design used Taylor-series linearization with strata `VARSTR`, PSU `VARPSU`, and weight `PERWT20F`. Payer interaction (fragmentation×insurance) was tested for MRI/CT-only; within-payer contrasts (Medium vs Low; High vs Low) were estimated. Unstable figure cells (CV>30% or unweighted n<50) were suppressed. Imaging flags missingness was set to FALSE/0 when no OB imaging records were observed; no imputation for covariates. Software: R 4.4.1; `survey` 4.4.2; `emmeans` 1.11.2; `ggplot2` 3.5.2.

## 3 Results

### 3.1 Cohort characteristics

We identified a spine cohort of 1,988 adults (weighted ∼22.8 million). The weighted probability of MRI/CT-only imaging in the spine cohort was ∼13.6%, with higher rates among those with multimorbidity (≥2 conditions) and increasing fragmentation levels.

**Table 2:** Spine cohort overview.

| Metric | Value |
| --- | --- |
| Unweighted N (persons) | 1,988 |
| Weighted population (persons) | $\sim 22,767,761$ |
| MRI/CT-only prevalence (weighted) | $\sim 13.6\%$ |

### 3.2 MRI/CT-only imaging by fragmentation and multimorbidity

Compared with Low fragmentation, the odds of advanced imaging were higher in Medium (OR 2.47, 95% CI 1.54–3.97) and High (OR 3.44, 95% CI 2.09–5.66) groups after adjustment.

**Figure 1.**
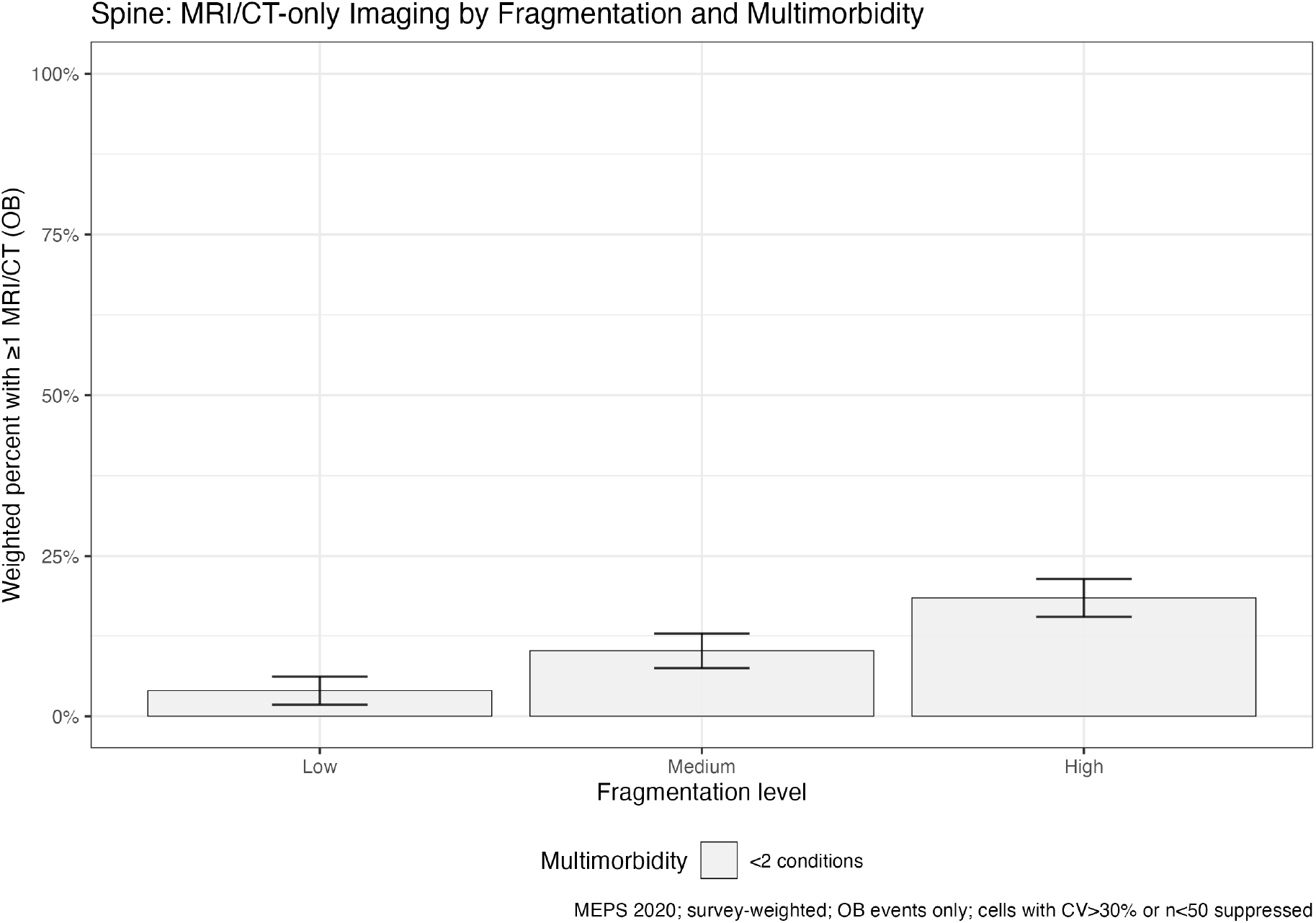
Survey-weighted probability (95% CI) of ≥1 MRI/CT in OB care by fragmentation level (Low →Medium →High) and multimorbidity (mcc 2plus). Cells with CV>30% or unweighted n<50 suppressed; unweighted cell Ns available in the appendix CSV.

### 3.3 Radiology expenditures by fragmentation and insurance

In the two-part model, any radiology spend was more likely in Medium (OR 1.71, 95% CI 1.10–2.67) and High (OR 2.00, 95% CI 1.27–3.15) vs Low fragmentation. Among those with positive spend, High vs Low showed a directionally higher expenditure (exp(beta) 1.56, 95% CI 0.96–2.55), though imprecise.

**Figure 2.**
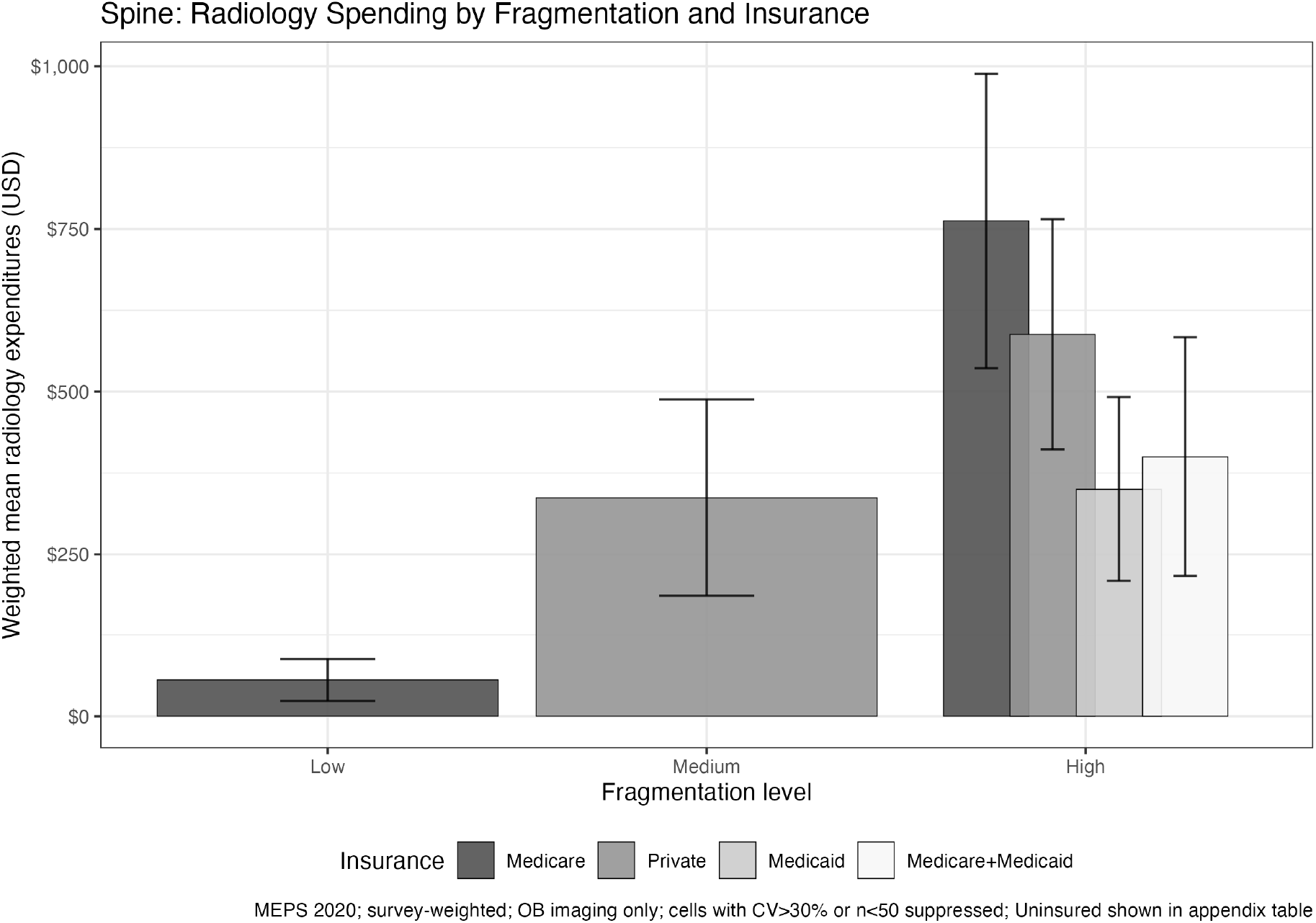
Survey-weighted mean radiology spending (USD; 95% CI) by fragmentation level (Low →Medium →High) and insurance. Cells with CV>30% or unweighted n<50 suppressed; unweighted cell Ns available in the appendix CSV.

### 3.3 Payer interaction for MRI/CT-only

Within-payer contrasts indicated the strongest association in private insurance (High vs Low OR 3.74, 95% CI 1.16–12.08). Medicare showed a similar trend (High vs Low OR 3.07, 95% CI 0.89–10.55) but was borderline; Medicaid and dual-eligible strata were imprecise.

**Table 3:** Payer-specific contrasts for MRI/CT-only (reference=Low)

| Insurance | Contrast | OR | 95% CI |
| --- | --- | --- | --- |
| Private | High vs Low | 3.74 | 1.16–12.08 |
| Private | Medium vs Low | 2.03 | 0.54–7.57 |
| Medicare | High vs Low | 3.07 | 0.89–10.55 |
| Medicare | Medium vs Low | 1.59 | 0.45–5.67 |

Global interaction (fragmentation×insurance) Wald F=72.53, p<0.001 (survey-weighted).

**Table 4:**
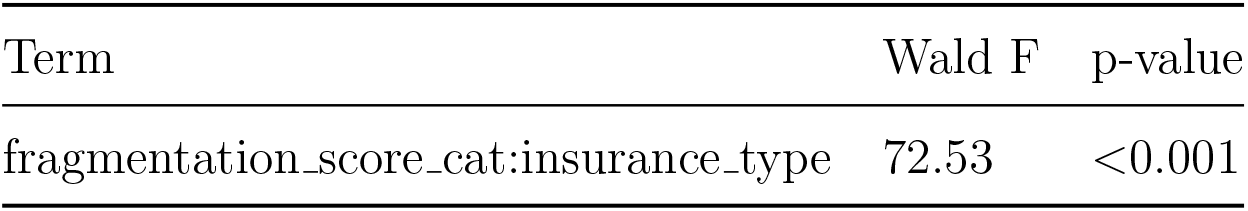
Global interaction test for MRI/CT-only.

| Term | Wald F | p-value |
| --- | --- | --- |
| fragmentation_score_cat:insurance_type | 72.53 | <0.001 |

## 4 Discussion

In a nationally representative cohort of U.S. adults with spine-related conditions, higher care fragmentation was associated with greater outpatient MRI/CT use and higher radiology spending. Associations were monotonic across fragmentation categories and appeared strongest among privately insured individuals, with similar (but less precise) trends in Medicare. These findings are consistent with concerns that fragmented ambulatory care can increase imaging intensity without necessarily improving downstream outcomes and complement operating-room evidence in which intraoperative monitoring benefits are uncertain [1]. Strengths include the use of survey-weighted methods, a prespecified fragmentation definition, and transparent suppression of unstable figure cells.

This analysis has limitations. First, MEPS public-use files lack procedural identifiers and intraoperative monitoring details; clinical severity and red-flag status are not directly observed. Second, results reflect office-based imaging only; emergency/outpatient department imaging is not included. Third, diagnosis-based cohorting may misclassify some conditions, and residual confounding may persist despite adjustment. Finally, 2020 utilization patterns occurred during the COVID-19 pandemic, which may affect health-seeking behavior and care patterns.

Taken together, the results support efforts to improve coordination in ambulatory spine care and to align imaging more closely with appropriateness frameworks—avoiding routine imaging for uncomplicated back pain while ensuring timely MRI for red-flag presentations [2, 4, 5, 3]. Payer differences suggest that benefit design, referral pathways, and authorization processes may shape imaging decisions in fragmented care.

## 5 Conclusions

Among U.S. adults with spine-related conditions, higher care fragmentation is associated with greater outpatient MRI/CT use and higher radiology spending. Effects appear most pronounced among privately insured individuals. Future work should extend beyond office-based settings and evaluate longitudinal episodes linking imaging to interventions and outcomes.

## Data Availability

All data produced in the present study are available upon reasonable request to the authors

## Data and code availability

The analysis uses the Medical Expenditure Panel Survey (MEPS) public-use files for 2020, which are de-identified and publicly available from the Agency for Healthcare Research and Quality (AHRQ) [6]. This study involved secondary analysis of public-use data and did not constitute human subjects research.

All analysis code (data processing, modeling, and figure generation) and a reproducible snapshot of derived outputs will be deposited in an open-access repository (e.g., Zenodo/OSF) under a versioned release and persistent DOI at the time of publication. The repository will include software environment details (R 4.4.1; `survey` 4.4.2; `emmeans` 1.11.2; `ggplot2` 3.5.2) and instructions to reproduce figures and tables from public MEPS inputs.

Supplementary materials provide:

- Table S1: Fragmentation category definitions (cutpoints).
- Tables S2–S6: Full model estimates for primary and secondary outcomes.
- Table S7: Listing of suppressed figure cells (high relative standard error or small unweighted cell counts).

## Supplementary material

**Table S1. Fragmentation cutpoints**

**Table 5:**
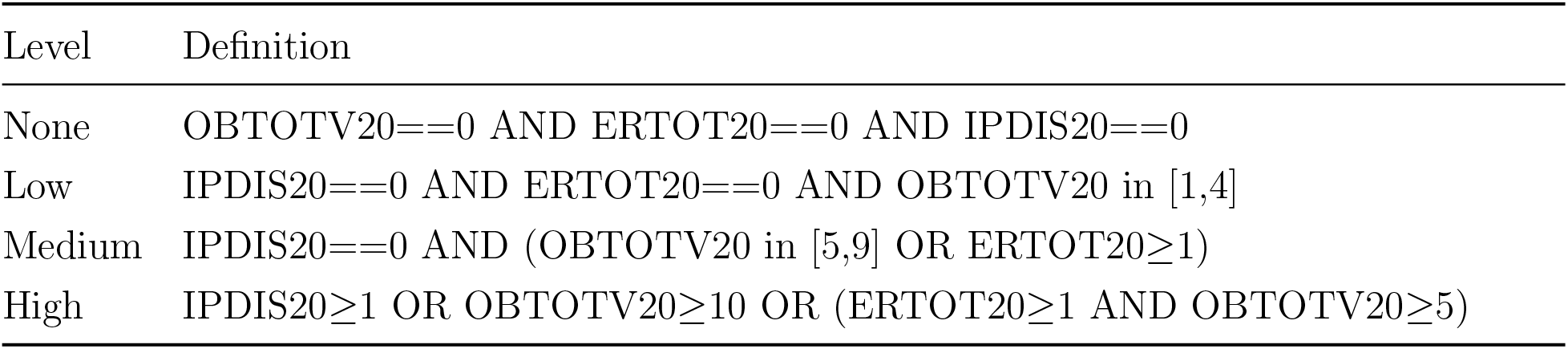
Table S1. Fragmentation cutpoints.

**Table S2. Full mod/CT-only (odds ratios)**

**Table 6:** Full model: MRI/CT-only (odds ratios)

| Term | OR | CI lower | CI upper | *p*-value |
| --- | --- | --- | --- | --- |
| (Intercept) | 0.01 | 0.00 | 0.07 | $1.37 \times 10^{-6}$ |
| fragmentation_score_catMedium | 2.10 | 1.13 | 3.87 | 0.02 |
| fragmentation_score_catHigh | 3.48 | 1.81 | 6.69 | $2.57 \times 10^{-4}$ |
| age_group35-49 | 1.67 | 0.72 | 3.85 | 0.23 |
| age_group50-64 | 2.38 | 1.03 | 5.49 | 0.04 |
| age_group65+ | 3.37 | 1.20 | 9.43 | 0.02 |
| female | 1.13 | 0.82 | 1.57 | 0.45 |
| race_ethnicityBlack | 1.51 | 0.45 | 5.05 | 0.50 |
| race_ethnicityHispanic | 1.22 | 0.41 | 3.60 | 0.72 |
| race_ethnicityOther | 2.19 | 0.67 | 7.18 | 0.20 |
| race_ethnicityWhite | 1.23 | 0.48 | 3.20 | 0.66 |
| poverty_statusLow Income | 0.55 | 0.32 | 0.95 | 0.03 |
| poverty_statusMiddle Income | 1.07 | 0.72 | 1.59 | 0.73 |
| poverty_statusNear Poor | 0.93 | 0.44 | 1.97 | 0.85 |
| poverty_statusPoor | 1.27 | 0.73 | 2.22 | 0.39 |
| insurance_typeMedicare | 0.66 | 0.26 | 1.67 | 0.38 |
| insurance_typeMedicare+Medicaid | 0.43 | 0.17 | 1.05 | 0.07 |
| insurance_typePrivate | 0.84 | 0.39 | 1.82 | 0.67 |
| insurance_typeUninsured/Other | 1.11 | 0.37 | 3.36 | 0.85 |
| OBTOTV20 | 1.01 | 1.01 | 1.02 | $6.14 \times 10^{-5}$ |
| mcc_2plus | 1.60 | 0.53 | 4.80 | 0.40 |

**Table S3. Full model: Any advanced imaging (odds ratios)**

**Table 7:** Full model: Any advanced imaging (odds ratios)

| Term | OR | CI lower | CI upper | *p*-value |
| --- | --- | --- | --- | --- |
| (Intercept) | 0.01 | 0.00 | 0.07 | $2.96 \times 10^{-7}$ |
| fragmentation_score_catMedium | 2.47 | 1.54 | 3.97 | $2.58 \times 10^{-4}$ |
| fragmentation_score_catHigh | 3.44 | 2.09 | 5.66 | $2.99 \times 10^{-6}$ |
| age_group35-49 | 0.98 | 0.55 | 1.75 | 0.94 |
| age_group50-64 | 1.41 | 0.83 | 2.38 | 0.21 |
| age_group65+ | 1.64 | 0.73 | 3.68 | 0.23 |
| female | 1.75 | 1.30 | 2.35 | $3.16 \times 10^{-4}$ |
| race_ethnicityBlack | 1.55 | 0.54 | 4.43 | 0.42 |
| race_ethnicityHispanic | 1.80 | 0.67 | 4.82 | 0.25 |
| race_ethnicityOther | 2.43 | 0.85 | 6.92 | 0.10 |
| race_ethnicityWhite | 1.41 | 0.56 | 3.50 | 0.47 |
| poverty_statusLow Income | 0.50 | 0.33 | 0.77 | 0.00 |
| poverty_statusMiddle Income | 0.97 | 0.70 | 1.34 | 0.86 |
| poverty_statusNear Poor | 1.01 | 0.50 | 2.04 | 0.98 |
| poverty_statusPoor | 0.94 | 0.61 | 1.44 | 0.77 |
| insurance_typeMedicare | 0.77 | 0.37 | 1.59 | 0.48 |
| insurance_typeMedicare+Medicaid | 0.67 | 0.34 | 1.30 | 0.24 |
| insurance_typePrivate | 0.69 | 0.41 | 1.16 | 0.16 |
| insurance_typeUninsured/Other | 0.72 | 0.30 | 1.75 | 0.47 |
| OBTOTV20 | 1.01 | 1.01 | 1.02 | $1.25 \times 10^{-4}$ |
| mcc_2plus | 3.28 | 1.10 | 9.74 | 0.03 |

**Table S4. Two-part model: Any radiology spend (odds ratios)**

**Table 8:** Two-part model: Any radiology spend (odds ratios)

| Term | OR | CI lower | CI upper | *p*-value |
| --- | --- | --- | --- | --- |
| (Intercept) | 0.10 | 0.03 | 0.35 | $3.85 \times 10^{-4}$ |
| fragmentation_score_catMedium | 1.71 | 1.10 | 2.67 | 0.02 |
| fragmentation_score_catHigh | 2.00 | 1.27 | 3.15 | 0.00 |
| age_group35-49 | 0.74 | 0.45 | 1.22 | 0.24 |
| age_group50-64 | 1.29 | 0.80 | 2.06 | 0.30 |
| age_group65+ | 1.81 | 0.86 | 3.80 | 0.12 |
| female | 1.71 | 1.35 | 2.18 | $2.34 \times 10^{-5}$ |
| race_ethnicityBlack | 1.60 | 0.73 | 3.48 | 0.24 |
| race_ethnicityHispanic | 1.40 | 0.63 | 3.11 | 0.41 |
| race_ethnicityOther | 2.00 | 0.78 | 5.17 | 0.15 |
| race_ethnicityWhite | 1.34 | 0.63 | 2.86 | 0.45 |
| poverty_statusLow Income | 0.90 | 0.62 | 1.30 | 0.56 |
| poverty_statusMiddle Income | 1.05 | 0.79 | 1.40 | 0.72 |
| poverty_statusNear Poor | 1.17 | 0.61 | 2.25 | 0.63 |
| poverty_statusPoor | 0.92 | 0.63 | 1.34 | 0.67 |
| insurance_typeMedicare | 0.71 | 0.39 | 1.31 | 0.28 |
| insurance_typeMedicare+Medicaid | 0.46 | 0.26 | 0.81 | 0.01 |
| insurance_typePrivate | 0.81 | 0.54 | 1.20 | 0.29 |
| insurance_typeUninsured/Other | 0.71 | 0.37 | 1.34 | 0.29 |
| OBTOTV20 | 1.02 | 1.01 | 1.03 | $2.32 \times 10^{-4}$ |
| mcc_2plus | 1.73 | 0.89 | 3.34 | 0.11 |

**Table S5. Two-part model: Positive radiology spend (exp(*β*))**

**Table 9:** Two-part model: Positive radiology spend (exp(*β*))

| Term | $\exp(\beta)$ | CI lower | CI upper | $p$-value |
| --- | --- | --- | --- | --- |
| (Intercept) | 504.24 | 147.57 | 1 722.99 | $1.91 \times 10^{-17}$ |
| fragmentation_score_catMedium | 1.18 | 0.69 | 2.02 | 0.54 |
| fragmentation_score_catHigh | 1.56 | 0.96 | 2.55 | 0.07 |
| age_group35-49 | 0.66 | 0.40 | 1.07 | 0.09 |
| age_group50-64 | 0.74 | 0.49 | 1.13 | 0.16 |
| age_group65+ | 0.60 | 0.32 | 1.13 | 0.12 |
| female | 1.06 | 0.76 | 1.49 | 0.72 |
| race_ethnicityBlack | 0.48 | 0.21 | 1.11 | 0.09 |
| race_ethnicityHispanic | 1.40 | 0.57 | 3.44 | 0.46 |
| race_ethnicityOther | 0.88 | 0.36 | 2.14 | 0.77 |
| race_ethnicityWhite | 0.94 | 0.42 | 2.10 | 0.88 |
| poverty_statusLow Income | 0.76 | 0.51 | 1.12 | 0.17 |
| poverty_statusMiddle Income | 0.90 | 0.65 | 1.26 | 0.55 |
| poverty_statusNear Poor | 1.45 | 0.64 | 3.28 | 0.37 |
| poverty_statusPoor | 1.66 | 0.76 | 3.63 | 0.20 |
| insurance_typeMedicare | 2.25 | 1.17 | 4.33 | 0.02 |
| insurance_typeMedicare+Medicaid | 1.16 | 0.66 | 2.04 | 0.61 |
| insurance_typePrivate | 1.96 | 1.16 | 3.31 | 0.01 |
| insurance_typeUninsured/Other | 1.14 | 0.59 | 2.21 | 0.70 |
| OBTOTV20 | 1.01 | 1.01 | 1.02 | $5.62 \times 10^{-7}$ |
| mcc_2plus | 0.81 | 0.45 | 1.49 | 0.51 |

**Table S6. Imaging-visit counts (rate ratios)**

**Table 10:** Imaging-visit counts (rate ratios)

| Term | Rate ratio | CI lower | CI upper | *p*-value |
| --- | --- | --- | --- | --- |
| (Intercept) | 0.14 | 0.04 | 0.43 | $7.28 \times 10^{-4}$ |
| fragmentation_score_catMedium | 2.48 | 1.61 | 3.84 | $6.80 \times 10^{-5}$ |
| fragmentation_score_catHigh | 4.09 | 2.79 | 6.00 | $2.78 \times 10^{-11}$ |
| age_group35-49 | 0.75 | 0.46 | 1.23 | 0.25 |
| age_group50-64 | 0.89 | 0.63 | 1.25 | 0.49 |
| age_group65+ | 0.54 | 0.18 | 1.60 | 0.27 |
| female | 1.59 | 1.14 | 2.23 | 0.01 |
| race_ethnicityBlack | 1.54 | 0.76 | 3.13 | 0.24 |
| race_ethnicityHispanic | 2.04 | 0.99 | 4.21 | 0.06 |
| race_ethnicityOther | 3.23 | 1.10 | 9.51 | 0.03 |
| race_ethnicityWhite | 1.83 | 0.92 | 3.62 | 0.09 |
| poverty_statusLow Income | 0.91 | 0.61 | 1.34 | 0.63 |
| poverty_statusMiddle Income | 1.00 | 0.67 | 1.49 | 1.00 |
| poverty_statusNear Poor | 1.15 | 0.58 | 2.27 | 0.69 |
| poverty_statusPoor | 1.05 | 0.64 | 1.72 | 0.84 |
| insurance_typeMedicare | 1.79 | 0.68 | 4.66 | 0.24 |
| insurance_typeMedicare+Medicaid | 1.01 | 0.58 | 1.75 | 0.98 |
| insurance_typePrivate | 1.08 | 0.70 | 1.65 | 0.73 |
| insurance_typeUninsured/Other | 0.99 | 0.48 | 2.03 | 0.98 |
| OBTOTV20 | 1.02 | 1.01 | 1.02 | $1.05 \times 10^{-11}$ |
| mcc_2plus | 0.97 | 0.43 | 2.17 | 0.94 |

**Table S7. Suppressed figure cells**

**Table 11:**
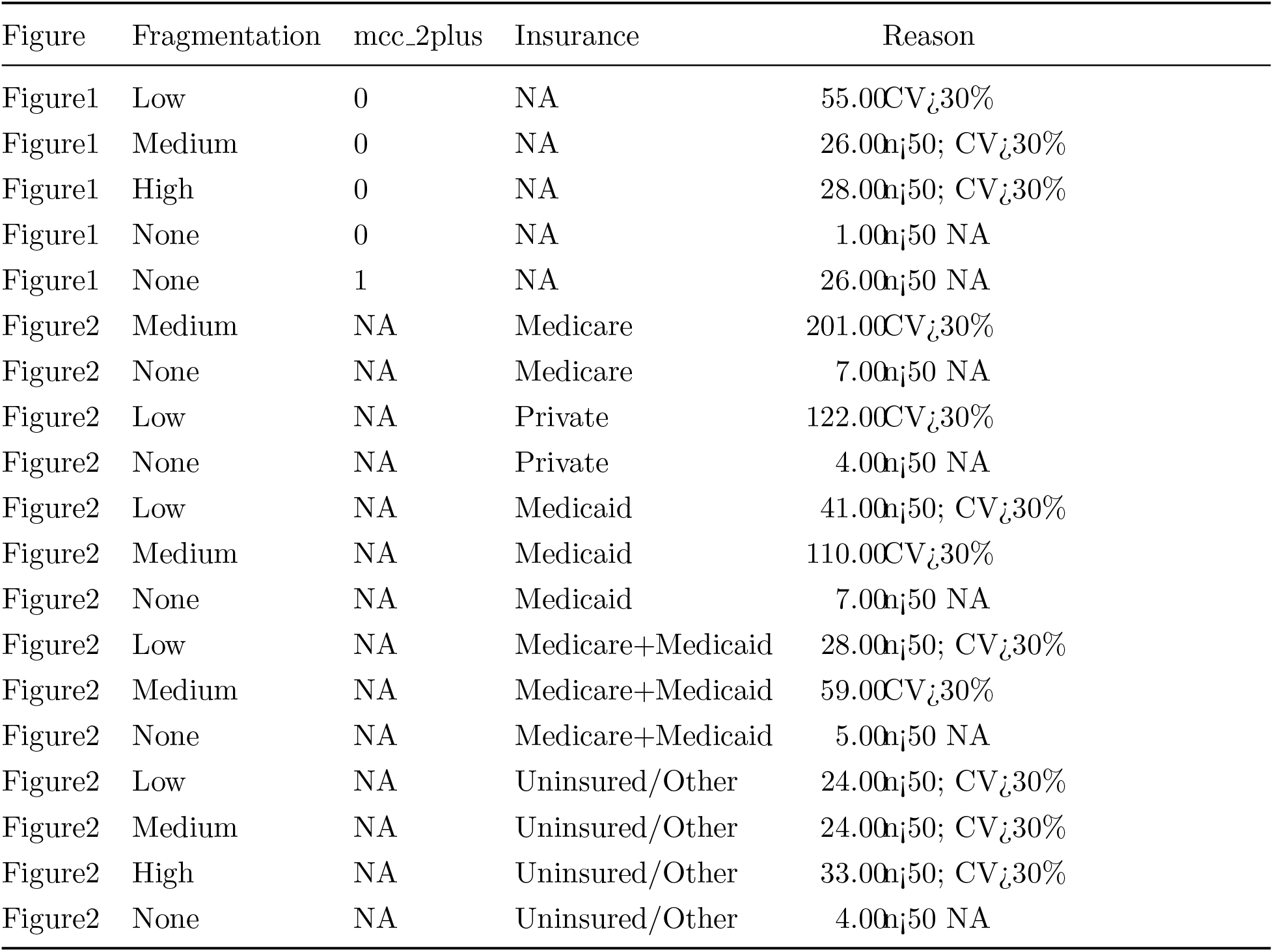
Suppressed figure cells.

